# Intertwined risk factors of mental health and cardiovascular diseases: A Cross-sectional survey in Godawari Municipality of Far-western Nepal

**DOI:** 10.1101/2025.03.11.25323720

**Authors:** Ramesh Ojha, Chiranjivi Adhikari, Hari Prasad Kaphle, Dikshya Adhikari

## Abstract

**Introduction:** Cardiovascular diseases (CVDs) are the leading global cause of death, whereas mental disorders account for one-third of all global disabilities. The significant relationship between these two giant co-morbidities, and their risk factors, is intertwined, and bidirectional. However, these relational factors are less explored in the Nepalese context. Hence, we aimed to determine the prevalence of mental health status and its associated factors among people with and without cardiovascular disease risk factors.

**Methods:** A community-based study was carried out during Sep-Nov 2024, among 390 people aged 30-69 years with and without cardiovascular disease risk factors in Godawari Municipality, Kailali. We Data was collected with face-to-face interviews using a structured questionnaire consisting of four sections: a) socio-demographic characteristics; b) CVD risk factors; c) mental health status; and d) anthropometric measurements. Data was collected using the KoboToolbox mobile application and imported into SPSS software for statistical analysis. We present categorical variables as frequency and percentage and continuous variables as median and quartiles. We applied univariate and multivariate logistic regression to determine factors associated with depression symptoms, anxiety, and stress. The result of logistic regression was presented as crude odds ratio, adjusted odds ratio (AOR), beta coefficients (β), and their 95% confidence interval, and p-values.

**Results:** The prevalences of depression, anxiety and stress symptoms were found 47.2%, 62.3% and 55.1%, respectively. From multivariate logistic regression analysis, depression symptoms were positively associated with females (β = 1.002, p < .001), and presence of 1 CVDs risk factor (β = 1.082, p = .016), 2 risk factors (β = 1.362, p = .006), and ≥ 3 risk factors (β = 1.720, p = .017). Anxiety symptoms were associated with exposure to secondhand smoking (β = 0.725, p = .024), and presence of 1 risk factor (β = 1.548, p < .001), 2 risk factors (β = 1.734, p < .001), and ≥ 3 risk factors (β = 1.852, p = .022). Dalit, Janajati and Madhesi (β = 0.735, p = .026), and presence of 1 risk factor (β = 1.811, p = .001), two risk factors (β = 2.054, p = .016), and ≥ 3 risk factors (β = 2.138, p < .001) were associated with stress symptoms.

**Conclusion:** The study revealed that nearly fifty percent of the prevalence of each of depression, anxiety, and stress symptoms among people with and without cardiovascular disease risk factors. Mental health screening is warranted among the people with two and above CVD risk factors.

## Introduction

Cardiovascular diseases (CVDs) are the global public health problem, causing 17.9 million deaths in 2019, and among them, more than three-quarters of deaths occur in low- and middle-income countries.^[1,2]^ Nepal is also facing an increasing burden of CVDs with a 3.8 case fatality rate and 1,104,474 disability-adjusted life years in 2019.^[3]^ Sudurpashchmin Province had a significantly higher percent (9.8%) of adults aged 40-69 with 30% or more CVD risk than almost all other provinces.^[4]^ Smoking and tobacco use (33.7%), alcohol consumption (20.8%), unhealthy diet (>90%), and hypertension (21%) were established risk factors for CVDs.^[4]^ These factors often lead to mental health morbidity and mortality. In 2019, 970 million people worldwide suffered from a mental disorder, the most frequent of which were anxiety and depression.^[5]^ According to NDHS 2022, the prevalence of symptoms of anxiety and/or depression in Far-western Nepal is 24.6% (urban 28.3% and rural 18.4%).^[6]^

There is a bidirectional relationship between mental health and CVDs.^[7]^ Mental health issues are more common in people with cardiovascular disease and its risk factors than in healthy people^[8]^ and severe mental illness has been linked to a two-fold increase in deaths from CVDs.^[9]^ Research on mental health is often given less emphasize, despite the fact that depression and anxiety are predicted to cost the global economy $1 trillion a year.^[10]^ The significant relationship of these two giant co-morbidities, and their risk factors is complex, intertwined, multi-faceted, and magnitudinous, and further implicates the healthcare delivery and outcomes. However, these relational factors are less explored in Far-western Nepal. Hence, the study aimed to assess the prevalence and risk factors associated with mental health status (depression, anxiety, and stress symptoms) among people with and without CVDs risk factors.

## Methods

### Study design, population, and setting

We conducted a cross-sectional study from September 1 through Nov 15, 2024, among participants aged 30-69 years with and without CVD risk factors in Godawari Municipality of Far-western Nepal.

### Sample size and sampling technique

The sample size was calculated by using the finite population correction (Cochrane) formula with a 5% margin of error and 95% confidence level and taking the prevalence of mental health status 50%. The calculated sample size was 384, but we ultimately enrolled 390 participants using a probability proportion to size sampling method. Out of 12 wards, one third wards were selected randomly using lottery method. From the selected wards, the required number of individuals were calculated based on probability proportionate to each ward and the households were selected purposively from each selected ward. If more than one eligible participant were present in the household, we used a lottery method to select the participant. The inclusion criteria for the study were people aged 30-69 years and who were permanent residents and living there for last 6 months in Godawari Municipality, Kailali. The exclusion criteria included: a) people who were ill and unable to communicate during the data collection period, b) people with known and diagnosed cardiac disease, using stent, c) pregnant women, and d) mentally retarded and bed-ridden patients.

### Data collection

We collected data with face-to-face interviews using a structured questionnaire consisting of four sections: a) socio-demographic characteristics; b) CVDs risk factors; c) mental health status; and d) anthropometric measurements. We collected data using electronic forms in the KoboToolbox, a free and open-source online data entry mobile application developed by Harvard Humanitarian initiatives.^[11]^ The participants were oriented about the purpose of the study before data collection. It took about 30–35 min for each participant to complete the interview.

### Variables and measurement

Mental health status: It was defined as the current state of depression, anxiety, and stress symptoms among the study participants. Nepali version of the validated Depression Anxiety and Stress Scale-21(DASS-21)^[12]^ was used to measure mental health status. The internal consistency (Cronbach’s alpha) of the DASS-21 Nepali version was 0.93 for DASS-depression; 0.79 for DASS-anxiety; and 0.91 for DASS-stress.^[13]^ DASS-21 is a widely used and validated tool for measuring mental health outcomes in many countries, including Nepal.^[14,15]^ There are 21 items on the DASS-21 scale (7 items each for depression, anxiety, and stress symptoms). Each participant is asked to score every item on a scale from 0 to 3, where 0 indicates “did not apply to me at all” and 3 indicates “apply to me at all”. The sum of each scale was multiplied by two to determine the overall scores for depression, anxiety, and stress symptoms. Depression, anxiety, and stress symptoms were further classified as binary (present/absent) if at least mild conditions were present.^[14]^

The independent variables assessed in this study included age (in years), gender (male/female), ethnicity (Dalit/Janajati/Brahmin/Chhetri), education status (illiterate/informal education/basic level/secondary level/bachelor’s or above), marital status (unmarried/married/wife), main occupation(unemployed/private employee/laborer/housekeeper/farmer/government job/business), and monthly family income (in NPR), smoking status (never/former smoker/current smoker), smokeless tobacco (never/former chewer/current chewer), frequency of consumption of smokeless tobacco (daily/sometimes), secondhand smoking (yes/no), fruits and vegetable intake (adequate/inadequate), body mass index status (underweight/normal/overweight/obese), blood pressure status (normal/hypertensive), body fat status (normal/excess), basal metabolic rate (lower/ideal), and hand grip strength (lower/ideal).

CVDs risk factors were defined as the presence of one or more risk factors in each participant, and these were assessed by counting the risk factors from current smoking and tobacco use, inadequate intake of fruits and vegetables, overweight/obesity, hypertension, and excess body fat. Thus, their presence was assigned a number between 0 to 5. Smoking and tobacco use ^[16]^, fruits and vegetables consumption ^[7]^, body mass index^[17]^, blood pressure^[18]^, body fat ^[19]^, basal metabolic rate ^[20]^, and hand-grip strength^[21]^ were defined using the protocol given by previous research. Fruit and vegetable consumption was further divided into low and high, taking the median as a cut-off point.

### Statistical analysis

KoboToolbox platform was used for data collection and entry.^[11]^ The data was cleaned and then exported to Statistical Package for Social Sciences (SPSS) version 25 for further analysis. Continuous variables were presented as median, and quartiles after checking for normal distribution whereas categorical variables were presented as frequencies and percentages. The normality of continuous variables was displayed visually (histogram) and numerically (Kolmogorov-Smirnov tests and Shapiro-Wilk test). Chi-square test (p-value <0.05 at 95% level of confidence) was used for testing for association between the dependent and independent variables (nominal and ordinal), and Mann Whitney U test statistic (p-value <0.05 at 95% level of confidence) was used for testing for association between the dependent and independent variables (continuous). We applied binary logistic regression to determine factors associated with depression symptoms, anxiety, and stress symptoms. In binary logistic regression analysis, we incorporated all variables with a chi-square test p-value below 0.25. Crude and adjusted odds ratio, beta coefficients (β) with 95% confidence intervals were calculated, and p-values < 0.05 were used to determine whether a variable was statistically significant. The association of independent and dependent variables was calculated by comparing association 0, 1, 2, and ≥3 risk factors of CVDs with mental health status. Before conducting the logistic regression, the Durbin-Watson test and variance inflation factor (VIF) with tolerance statistics were carried out to check the independence of errors and multicollinearity of the independent variables and revealed no multicollinearity. Parameters such as Negelkarke R squares, Cox and Snell R squares, the log likelihoods, and the Hosmer and Lemeshow test were carried out to determine the model’s goodness of fit and the variance of independent variables that described the mental health status.

### Ethical consideration

We obtained ethical approval from the Institutional review committee of Pokhara University (Reference Number: 96/2081/82-IRC, approval date: August 30, 2024). Formal permission was obtained from Godawari Municipality before conducting the study. We obtained written informed consent from each participant before enrolling them in the study. We ensured that participation in the study was voluntary and maintained confidentiality throughout the study.

## Results

### Socio-demographic, behavioral, and bio-physical characteristics of the study participants

Of 390 participants, two-third (66.9%) of them were females, with Brahmin/Chhetri as a major ethnic group (83.8%). Surprisingly, 11.8% were widower or widow. One in every five participants was illiterate. Among total participants, about one-third (32.1%) of them were overweight and obese. Nearly one third (30.3%) of respondents were hypertensive, whereas less than half (40.5%) of the participants had excess body fat. (table 1)

**Table 1:**
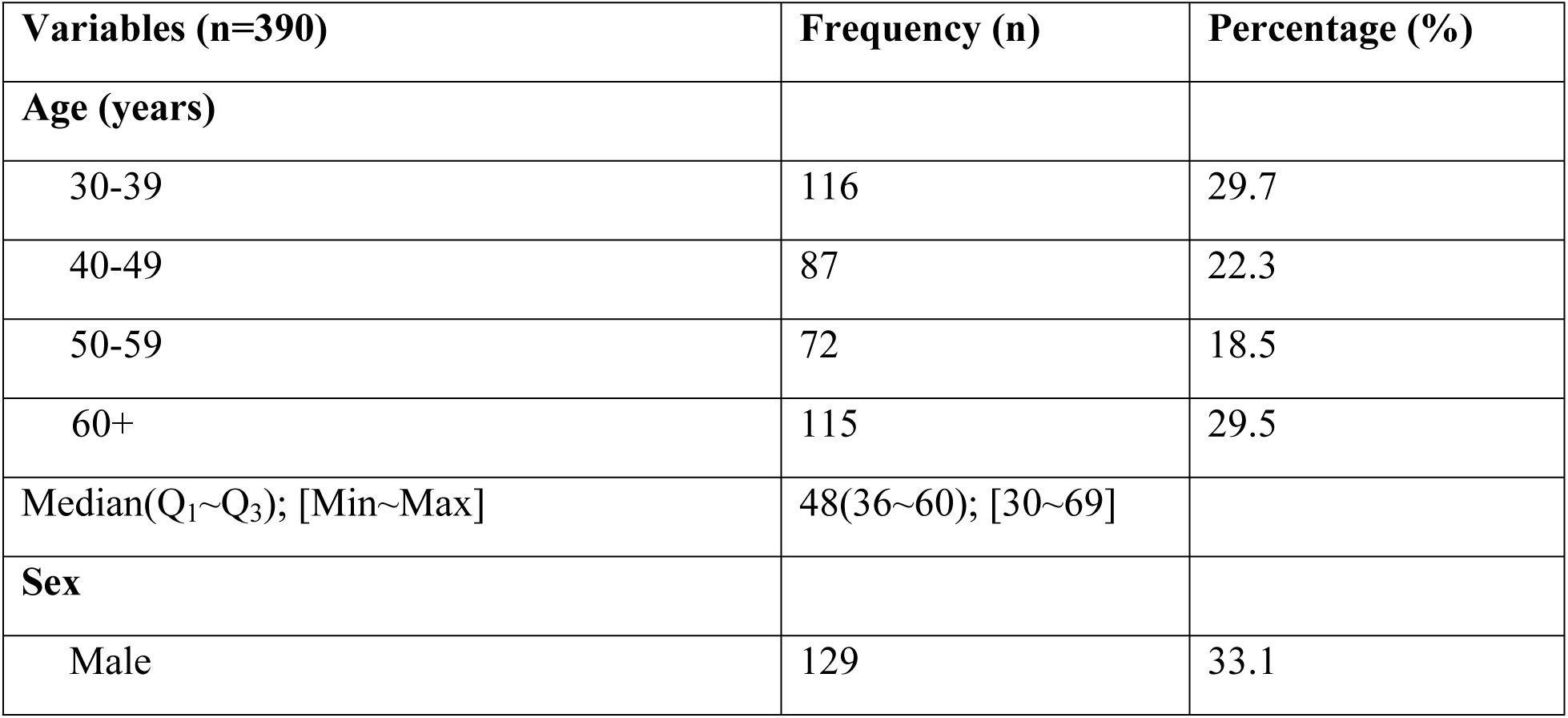
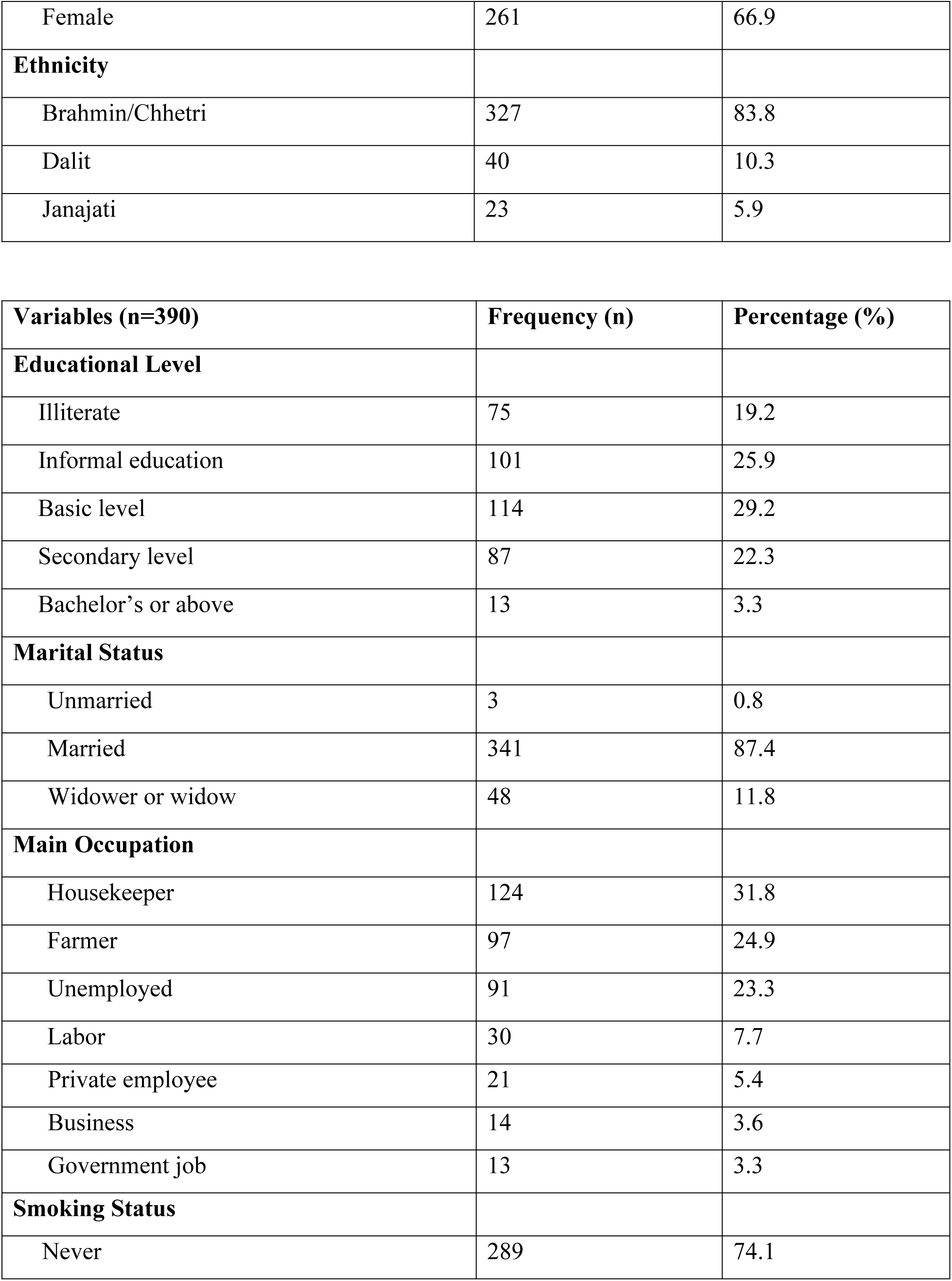
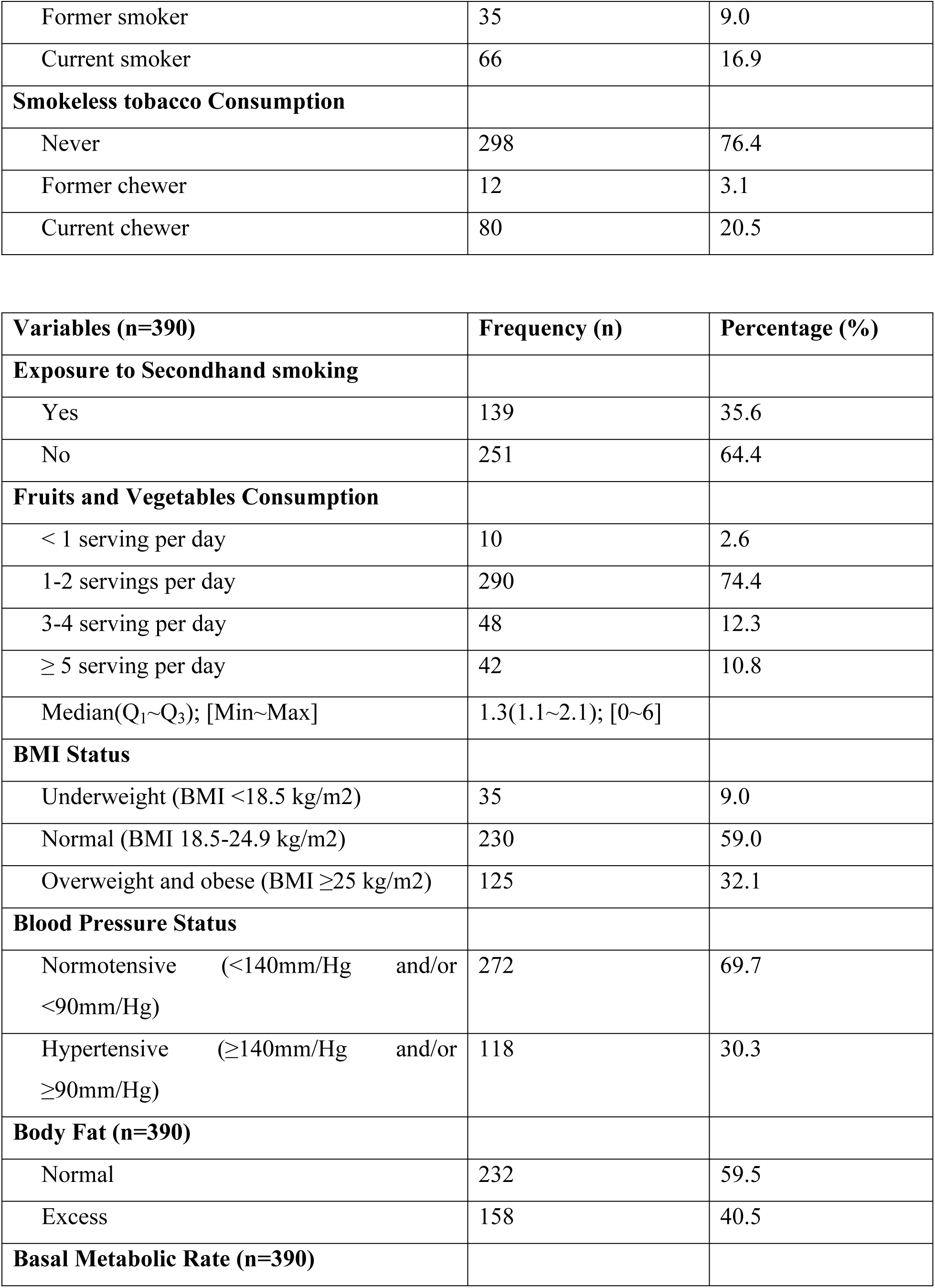
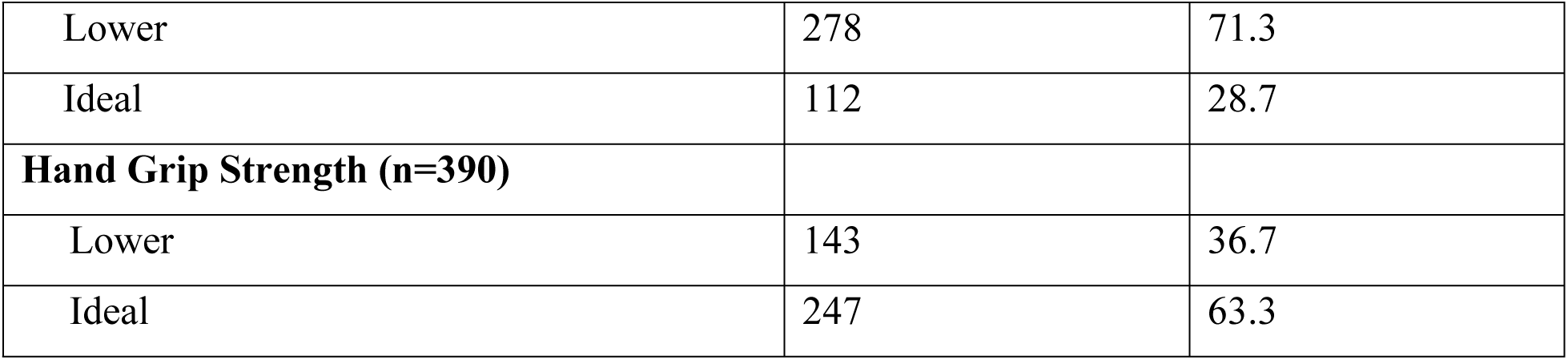
Socio-demographic, behavioral, and bio-physical characteristics of the study participants.

### Prevalence of number of CVDs risk factors among study participants

Furthermore, we assessed the prevalence of risk factors for CVDs risk factors by the number of factors across sexes and different age groups. The median risk factor (Q_1_∼Q_3_) was 2 (1∼3). Majority (88.2%) of them had one or more CVDs risk factors. (figure 1).

**Figure 1:**
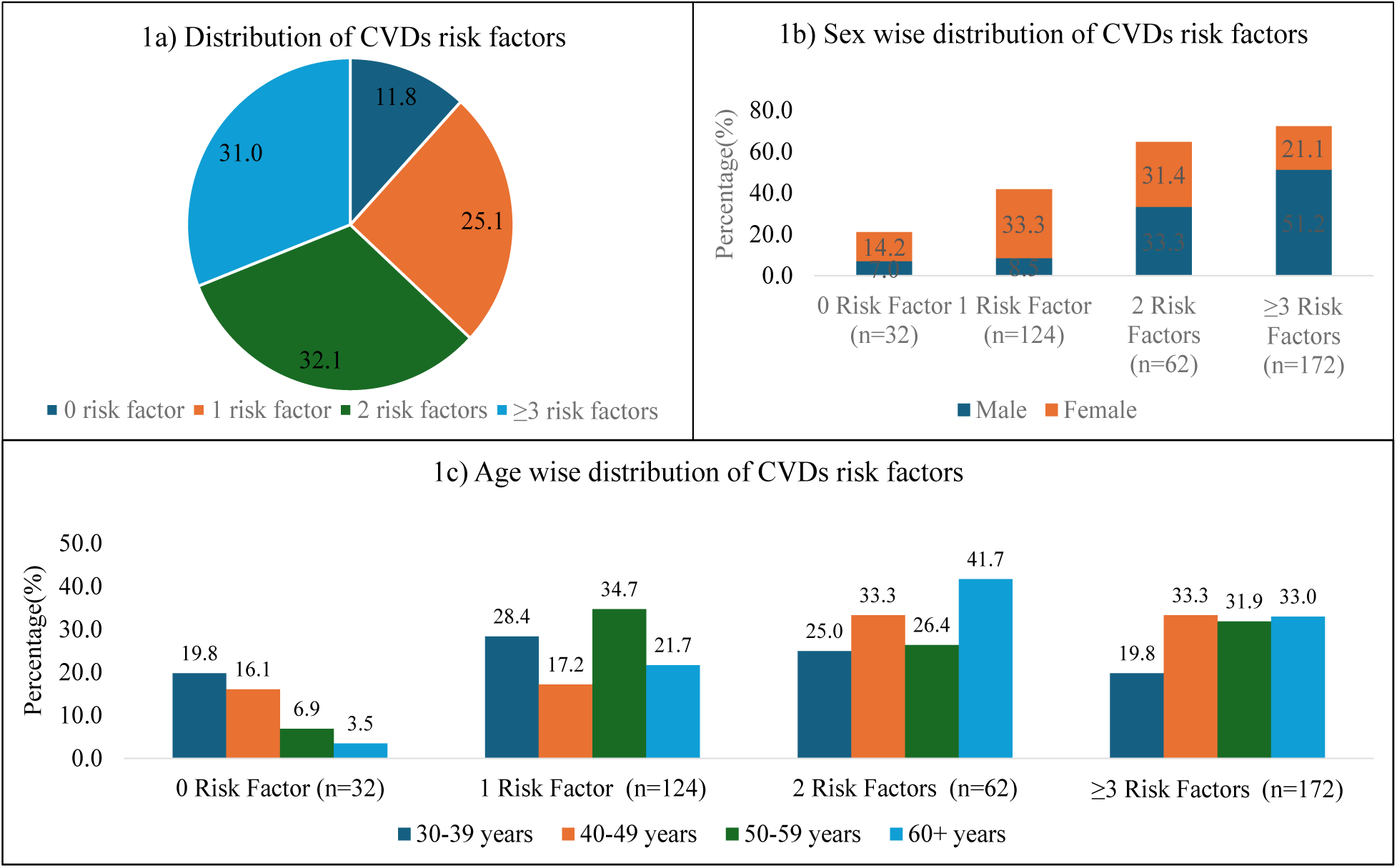
Prevalence of number of CVDs risk factors among study participants (n=390)

### Prevalence of mental health status (DASS 21 score)

The prevalence rates of depression, anxiety, and stress were found to be 47.2%, 62.3%, and 55.1% among people with and without CVDs risk factors (table 2).

**Table 2:** Prevalence of mental health status (DASS 21 score)

| **Variables (n=390)** | **Frequency (n)** | **Percentage (%)** |
| --- | --- | --- |
| **Depression (Score of Depression Subscale)** |  |  |
| Normal (0-9) | 206 | 52.8 |
| Mild (10-13) | 91 | 23.3 |
| Moderate (14-20) | 59 | 15.1 |
| Severe (21-27) | 32 | 8.2 |
| Extremely severe (28+) | 2 | 0.5 |
| Total with depression symptoms | 184 | 47.2 |
| Median(Q<sub>1</sub>~Q<sub>3</sub>); [Min~Max] | 14(16~22); [6~38] |  |
| **Anxiety (Score of Anxiety Subscale)** |  |  |
| Normal (0-7) | 147 | 37.7 |
| Mild (8-9) | 108 | 27.7 |
| Moderate (10-14) | 70 | 17.9 |
| Severe (15-19) | 60 | 15.4 |
| Extremely severe (20+) | 5 | 1.3 |
| Total with anxiety symptoms | 243 | 62.3 |
| Median(Q<sub>1</sub>~Q<sub>3</sub>); [Min~Max] | 9(7~14); [4~30] |  |
| **Stress (Score of Stress Subscale)** |  |  |
| Normal (0-14) | 175 | 44.9 |
| Mild (15-18) | 67 | 17.2 |
| Moderate (19-25) | 97 | 24.9 |
| Severe (26-33) | 49 | 12.6 |
| Extremely severe (34+) | 2 | 0.5 |
| Total with stress symptoms | 215 | 55.1 |
| Median(Q<sub>1</sub>~Q<sub>3</sub>); [Min~Max] | 9(8~13); [0~30] |  |

Participants constituting 6.9%, 11.0%, and 7.7% experienced only depression, anxiety, and stress symptoms, respectively, whereas 29.2% experienced all three mental health issues concurrently. (figure 2).

**Figure 2:**
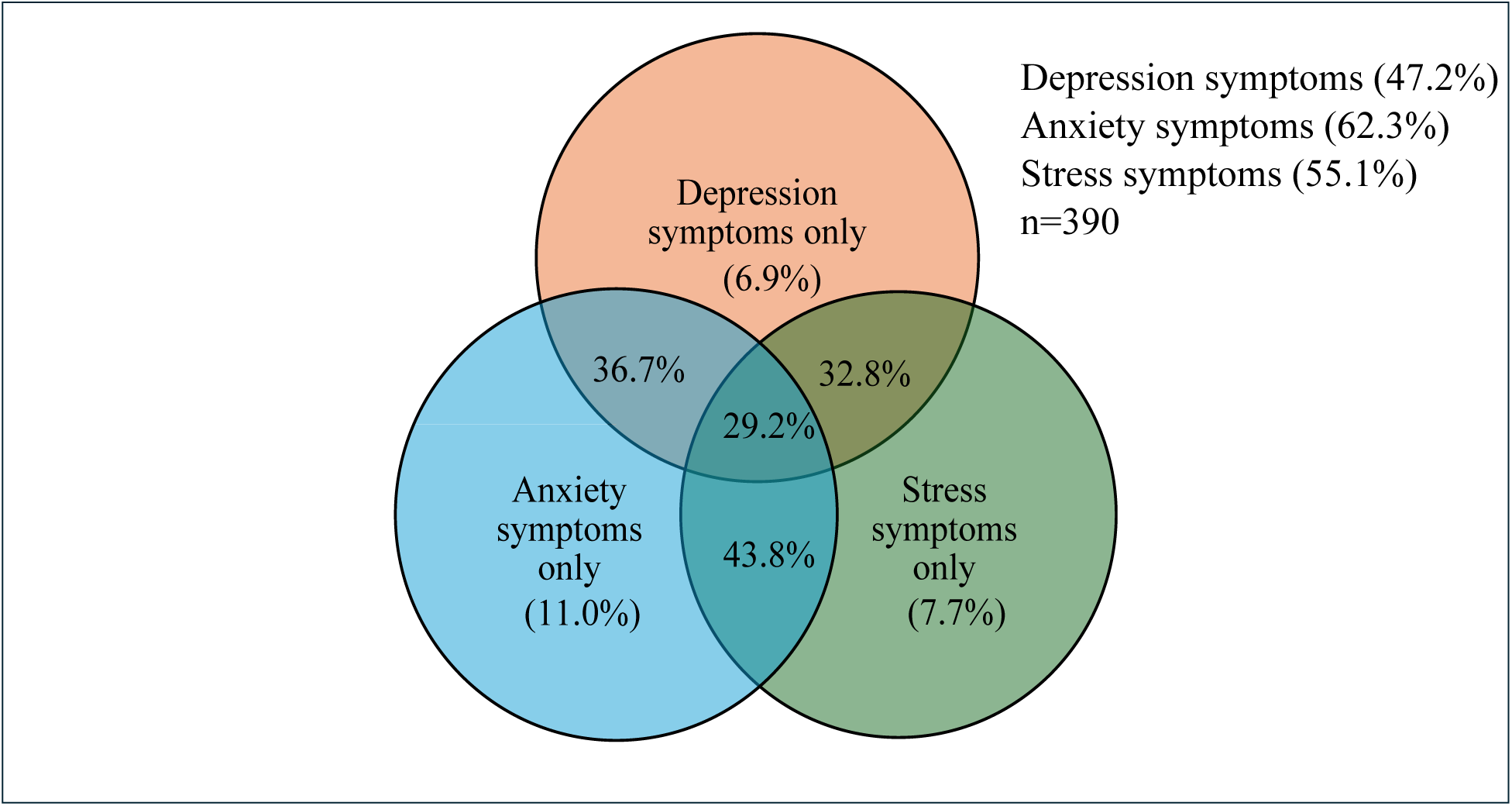
Prevalence of depression, anxiety and stress symptoms.

### Factors associated with depression symptoms

Multivariate analysis showed that sex, and the distribution of CVDs risk factors were statistically significantly associated with depression symptoms among people with and without CVDs risk factors. Female participants were two times (aOR: 2.725, 95% CI: 1.575 to 4.715) more likely to have depression symptoms in comparison with males. In comparison with zero risk factor, individuals with one risk factor (aOR: 2.952, 95% CI: 1.224 to 7.121), two risk factors (aOR: 3.904, 95% CI: 1.480 to 10.299) and three or more than three risk factors (aOR: 5.585, 95% CI: 1.357 to 22.991) were more likely to have depression symptoms. (table 3)

**Table 3:**
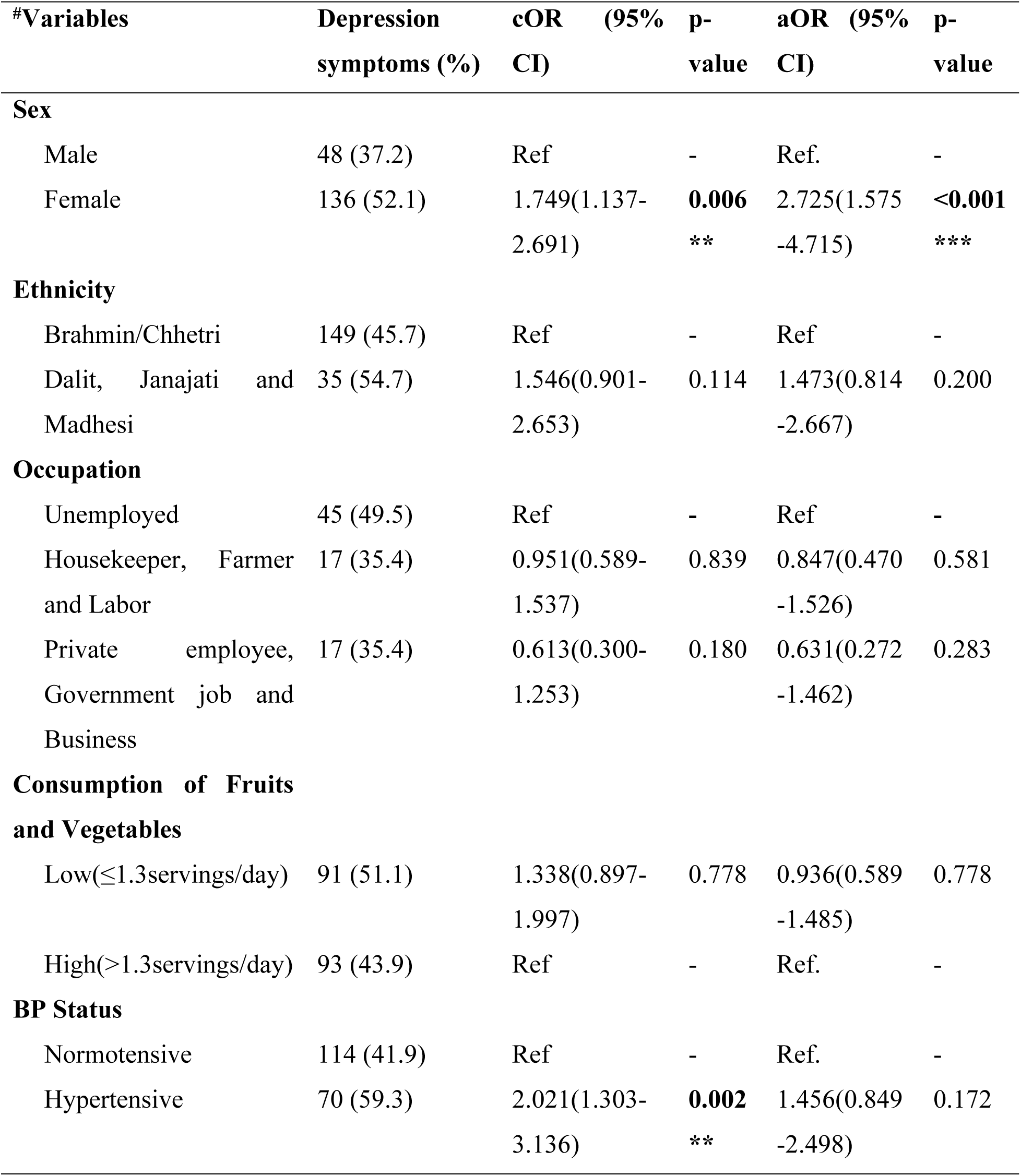
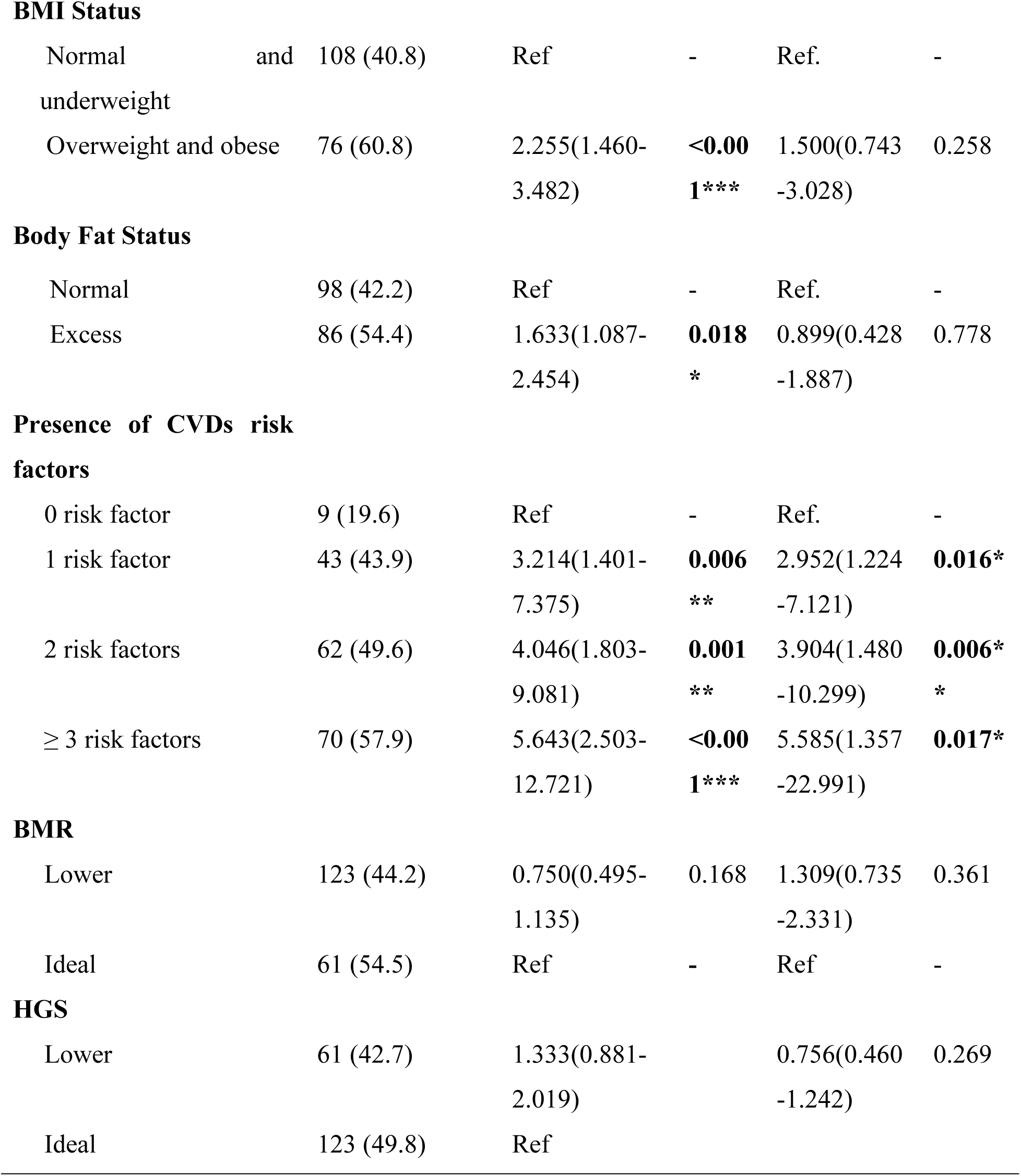
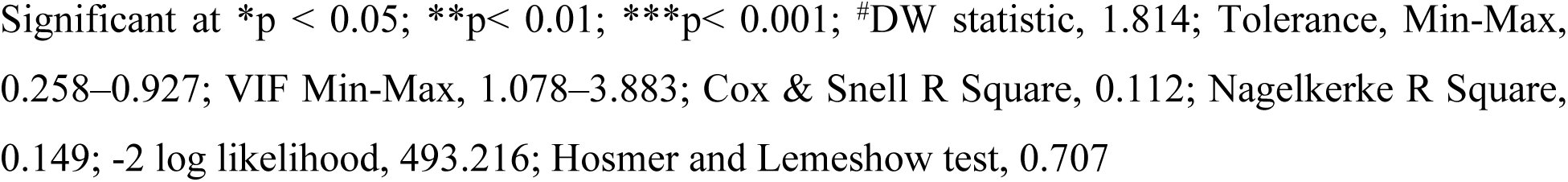
Factors associated with depression symptoms among people with and without CVDs risk factors.

### Factors associated with anxiety symptoms

Participants who were exposed to secondhand smoking were two-folds (aOR: 2.065, 95% CI: 1.100 to 3.877) more likely to have anxiety symptoms compared to those who were unexposed to secondhand smoking. In comparison with zero risk factor, participants with one risk factor (aOR: 4.703, 95% CI: 2.046 to 10.811), two risk factors (aOR: 5.664, 95% CI: 2.190 to 14.646) and with three or more than three risk factors (aOR: 6.374, 95% CI: 1.314 to 30.915) were more likely to have anxiety symptoms. (table 4)

**Table 4:** Factors associated with anxiety symptoms among people with and without CVDs risk factors.

| #Variables | Anxiety symptoms (%) | cOR (95% CI) | p-value | aOR (95% CI) | p-value |
| --- | --- | --- | --- | --- | --- |
| **Sex** |  |  |  |  |  |
| Male | 90 (69.8) | Ref. | - | Ref. | - |
| Female | 160 (61.3) | 1.629(1.039-2.553) | **0.033\*** | 1.338(0.673-2.657) | 0.406 |
| **Ethnicity** |  |  |  |  |  |
| Brahmin/Chhetri | 196 (60.1) | Ref | - | Ref | - |
| Dalit, Janajati and Madhesi | 47 (73.4) | 1.834(1.009-3.333) | **0.044\*** | 1.411(0.736-2.703) | 0.300 |
| **Smoking status** |  |  |  |  |  |
| Never or former smoker | 193 (59.6) | Ref | - | Ref | - |
| Current smoker | 50 (75.8) | 2.121(1.158-3.885) | **0.013\*** | 0.549(0.176-1.714) | 0.302 |

**Table 4** (continued)
| #Variables | Anxiety symptoms (%) | cOR (95% CI) | p-value | aOR (95% CI) | p-value |
| --- | --- | --- | --- | --- | --- |
| **Smokeless Tobacco** |  |  |  |  |  |
| Never or former consumer | 180 (58.1) | Ref |  | Ref |  |
| Current consumer | 63 (78.8) | 2.676(1.497-4.786) | **0.001\*\*** | 1.817(0.530-6.232) | 0.342 |
| **Secondhand smoke(ire)** |  |  |  |  |  |
| No | 138 (55.0) | Ref | - | Ref | - |
| Yes | 105 (75.5) | 2.529(1.597-4.005) | **<0.001\*\*\*** | 2.065(1.100-3.877) | **0.024\*** |
| **BP Status** |  |  |  |  |  |
| Normotensive | 156 (57.4) | Ref | - | Ref | - |
| Hypertensive | 87 (73.7) | 2.087(1.298-3.356) | **0.002\*\*** | 1.086(0.594-1.987) | 0.789 |
| **BMI Status** |  |  |  |  |  |
| Normal and underweight | 152 (57.4) | Ref | - | Ref | - |
| Overweight and obese | 91 (72.8) | 1.990(1.252-3.162) | **0.004\*\*** | 0.957(0.436-2.102) | 0.913 |
| **Body Fat Status** |  |  |  |  |  |
| Normal | 123 (53.0) | Ref | - | Ref | - |
| Excess | 120 (75.9) | 2.798(1.790-4.375) | **<0.001\*\*\*** | 1.499(0.676-3.323) | 0.319 |

**Table 4** (continued)
| #Variables | Anxiety symptoms (%) | cOR (95% CI) | p-value | aOR (95% CI) | p-value |
| --- | --- | --- | --- | --- | --- |
| **Presence of CVDs risk factors** |  |  |  |  |  |
| 0 risk factor | 10 (21.7) | Ref | - | Ref | - |
| 1 risk factor | 55 (56.1) | 4.605(2.056-10.312) | **0.002\*\*** | 4.703(2.046-10.811) | **<0.001\*\*\*** |
| 2 risk factors | 84 (67.2) | 7.376(3.334-16.315) | **<0.001\*\*\*** | 5.664(2.190-14.646) | **<0.001\*\*\*** |
| ≥ 3 risk factors | 94 (77.7) | 12.533(5.515-28.485) | **<0.001\*\*\*** | 6.374(1.314-30.915) | **0.022\*** |
| **BMR** |  |  |  |  |  |
| Lower | 166 (59.7) | 1.484(0.932-2.365) | 0.097 | 1.043(0.567-1.918) | 0.893 |
| Ideal | 77 (68.8) | Ref | - | Ref | - |
| **HGS** |  |  |  |  |  |
| Lower | 79 (55.2) | 0.625(0.410-0.953) | **0.026\*** | 0.644(0.390-1.063) | 0.085 |
| Ideal | 164 (66.4) | Ref | - | Ref | - |

Significant at \*p < 0.05; \*\*p< 0.01; \*\*\*p< 0.001; #DW statistic, 1.941; Tolerance, Min-Max, 0.243–0.935; VIF Min-Max, 1.070–4.112; Cox & Snell R Square, 0.143; Nagelkerke R Square, 0.194; -2 log likelihood, 456.736; Hosmer and Lemeshow test, 0.796

### Factors associated with stress symptoms among people with and without CVDs risk factors (n=390)

From multi-variate logistic regression, Dalit, Janajati, and Madhesi participants were 2 times (aOR: 2.085, 95% CI: 1.091 to 3.985) more likely to have stress symptoms compared to those Brahmin/Chhetri. In comparison with zero risk factor, participants with one risk factor (aOR: 6.119, 95% CI: 2.202 to 17.008), two risk factors (aOR: 7.800, 95% CI: 1.463 to 41.582) and participants with three or more than three risk factors (aOR: 8.484, 95% CI: 2.769 to 25.997) were more likely to have stress. (table 5)

**Table 5:**
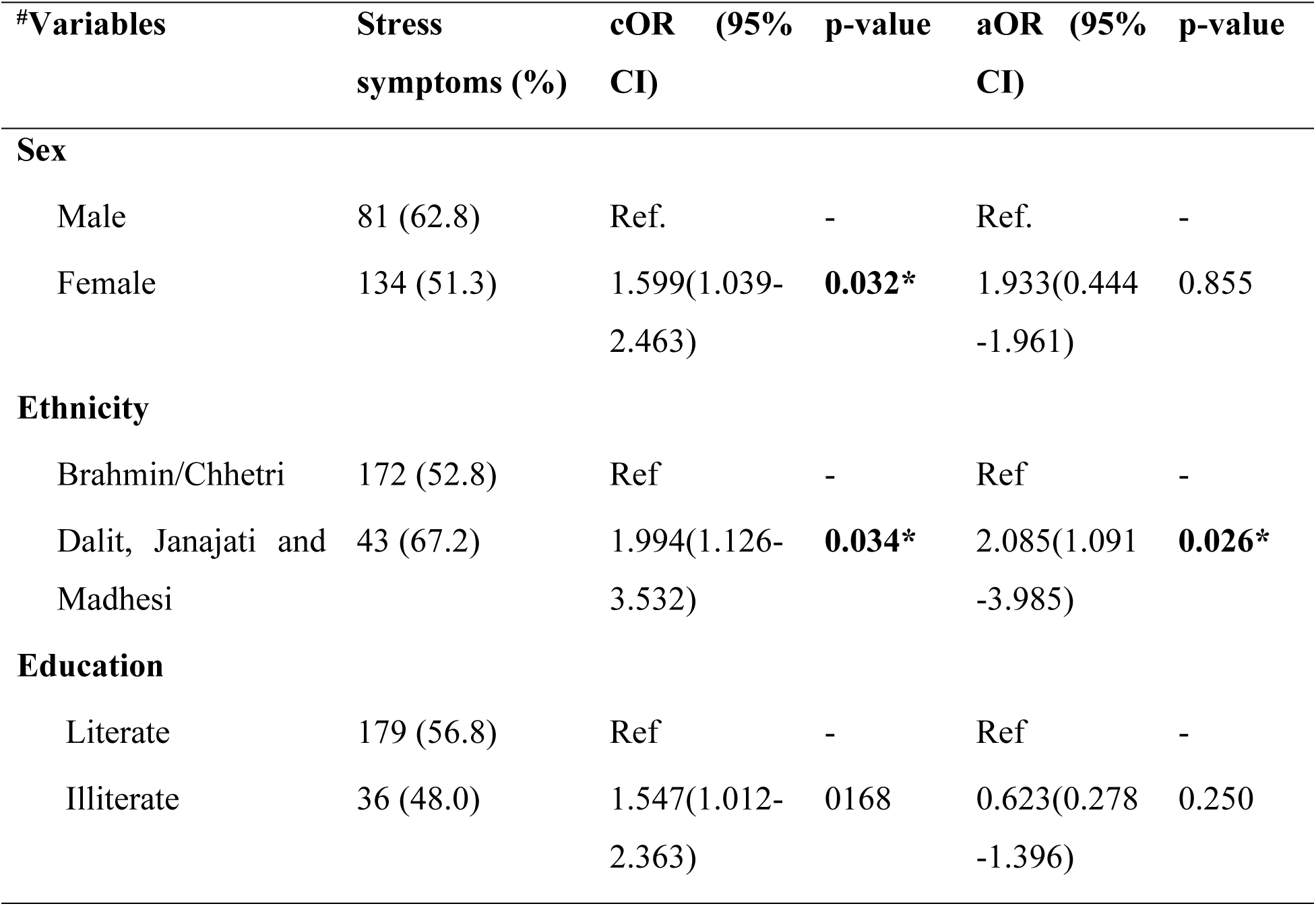
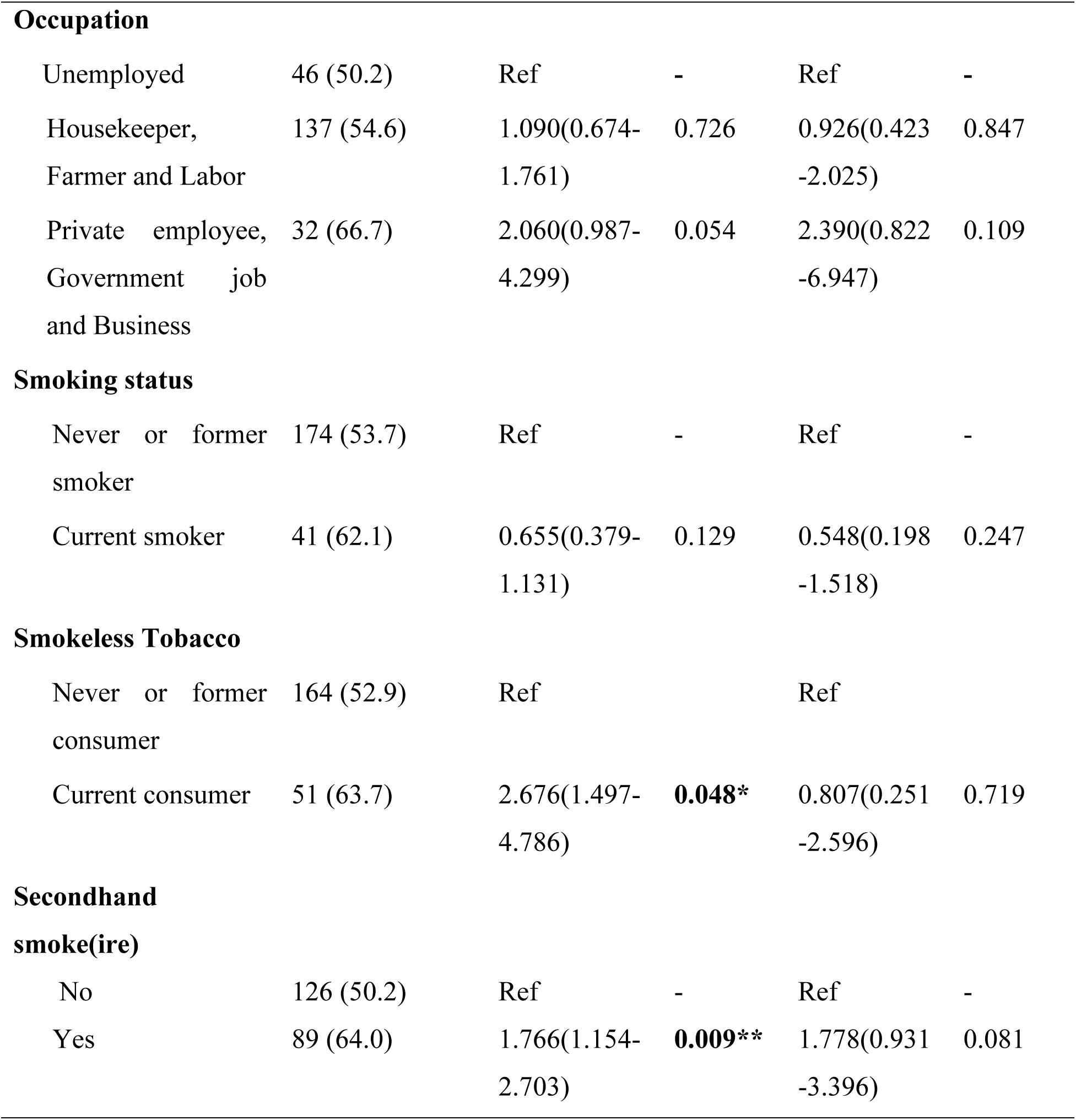
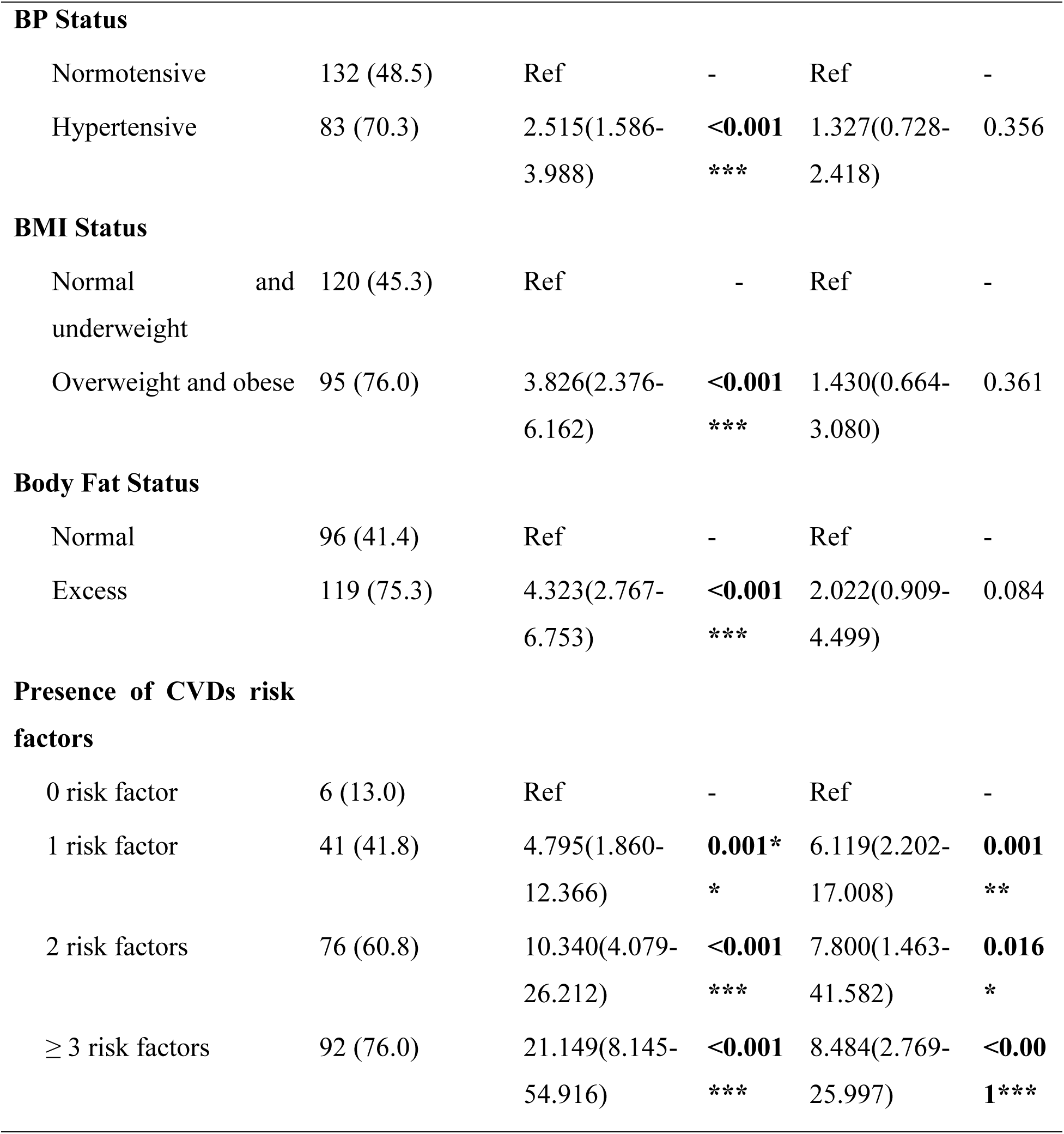
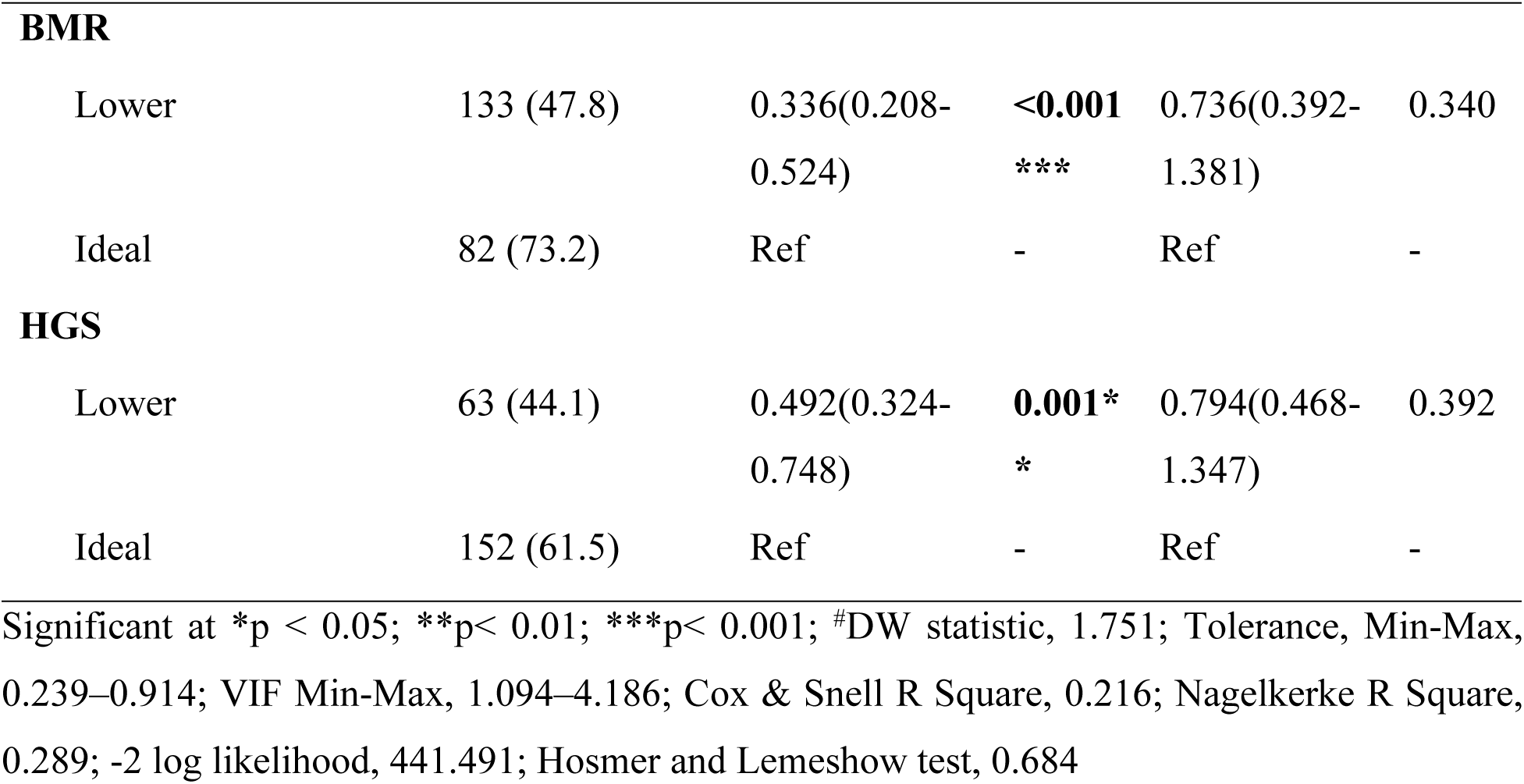
Factors associated with stress symptoms among people with and without CVDs risk factors.

### Predictors and their β-values (S.E.) of mental health status

Distribution of CVD risk factors were significantly associated with mental health status. As the number of risk factors increases, depression, anxiety, and stress symptoms also increase significantly. (table 6, figure 3)

**Figure 3:**
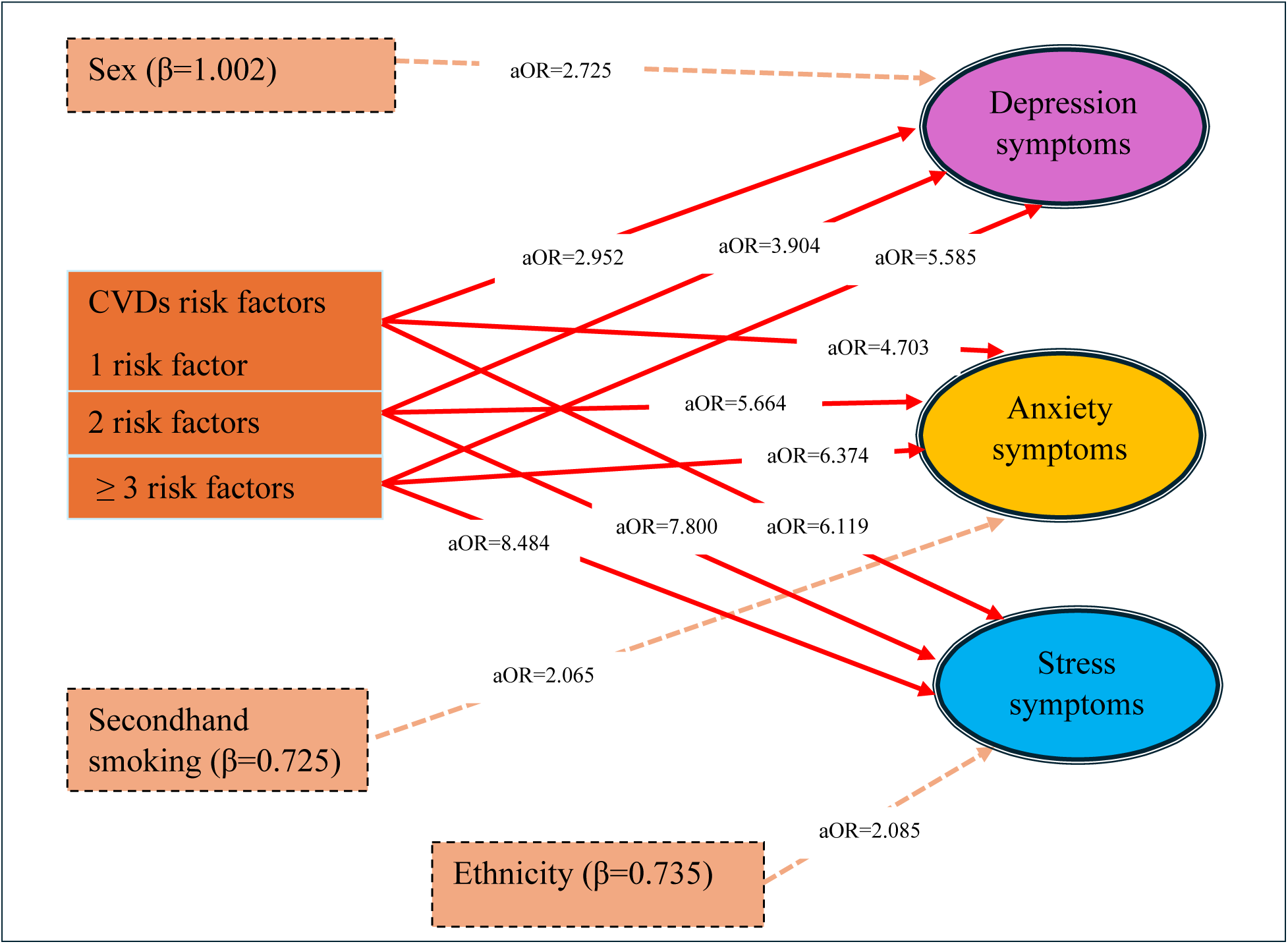
Predictors of depression, anxiety, and stress symptoms.

**Table 6:**
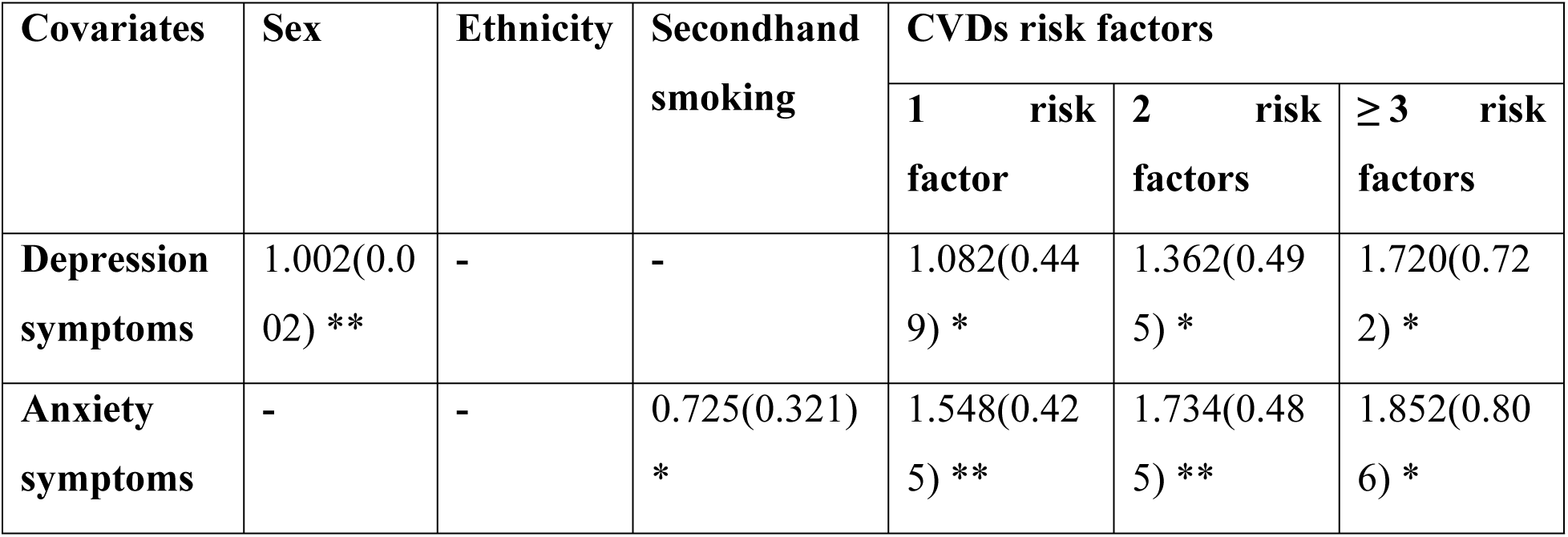
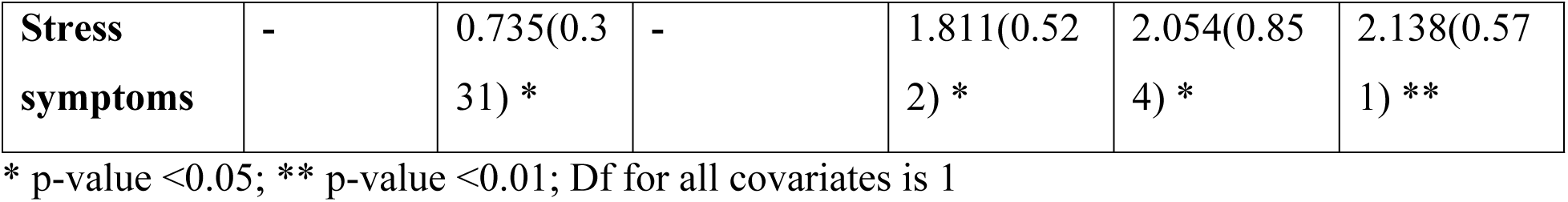
Predictors and their β-values (S.E.) of mental health status.

| Covariates | Sex | Ethnicity | Secondhand smoking | CVDs risk factors |  |  |
| --- | --- | --- | --- | --- | --- | --- |
|  |  |  |  | 1 risk factor | 2 risk factors | ≥ 3 risk factors |
| Depression symptoms | 1.002(0.002) ** | - | - | 1.082(0.449) * | 1.362(0.495) * | 1.720(0.722) * |
| Anxiety symptoms | - | - | 0.725(0.321) * | 1.548(0.425) ** | 1.734(0.485) ** | 1.852(0.806) * |
| **Stress symptoms** | - | 0.735(0.331) * | - | 1.811(0.522) * | 2.054(0.854) * | 2.138(0.571) ** |

\* p-value <0.05; \*\* p-value <0.01; Df for all covariates is 1

## Discussion

From this study, prevalence of depression, anxiety, and stress were found to be 47.2%, 62.3%, and 55.1%, respectively. Firstly, sex, and CVDs risk factors were revealed as risk factors of depression symptoms; Secondhand smoking, and CVDs risk factors of anxiety symptoms; and ethnicity, and CVDs risk factors of stress symptoms. Previous studies identified smoking habits, Blood pressure, and body fat were associated with depression, anxiety, and stress symptoms but these are found to be non-significant in this study. We further discuss the prevalence, risk factors, and non-significant factors.

The prevalence of depression, anxiety, and stress symptoms in this study is consistent with the findings reported by Thagunna and his colleagues among the Nepalese youth population, with 50.6% depression, 46.5% anxiety, and 56.2% stress.^[22]^ In contrary, the prevalence of depression and anxiety symptoms in this study was much higher than previously reported findings by NDHS 2022 among adults’ populations of Nepal. ^[6]^ This was due to the sample population taken in NDHS survey was age group 15-49 years, and more than half of the women and men interviewed were under age 30, but in this study the sample population was aged 30-69 years. The prevalence of stress symptoms was consistent with findings reported by the NCDs-STEPS Survey 2019 with 62.3% stress.^[4]^ Similarly, the findings of this study are consistent with the findings presented by the study among chronic disease patients in Karnatak, India.^[8]^ A study conducted by Basnet and his colleagues among residents of Nepal reported 35.1% depression, 31% anxiety and 22.5% stress, which was lower than this study.^[23]^ One convincible reason for this variation may be that CVDs risk factors are significantly associated with mental health status and people with these risk factors are at higher risk of developing poor mental health. The prevalence of depression, anxiety, and stress symptoms was higher than the findings reported by a study conducted among construction workers in Kavre district, Nepal,^[14]^ and among traffic police officers in Kathmandu.^[15]^ The findings of this study are higher than the study conducted among India^[24]^, Northwest Iran^[25]^, and Ghana.^[26]^ The possible reasons for the variation in the prevalence of mental health status may be due to differences in the study setting, study period, study population, study variables, socioeconomic status, sample size and sampling techniques, study tool and techniques, data analysis methods, and COVID-19 pandemic.

The findings of this study revealed that the distribution of number of CVDs risk factors was significantly associated with depression, anxiety, and stress symptoms. Findings of previous studies.^[27,28]^ showed that people with CVDs risk factors have a greater risk of mental health disorders compared to the general population The possible justification could be that prior studies reported that many CVDs risk factors, such as smoking habits, harmful use of alcohol, hypertension, diabetes, excess body fat, low intake of fruits and vegetables, and overweight and obese were more prevalent. Risk factors of CVDs were significantly associated with mental health status and people with these risk factors are at higher risk of developing poor mental health such as depression, anxiety, and stress.

In this study, females were nearly three times higher risk of having depression symptoms compared to those males. This finding was consistent with the previous studies including the 2022 Nepal Demographic and Health Survey (NDHS), have shown that females experience a higher prevalence of depression (5.4% (95%CI: 4.8, 6.1) among females and 1.7% (95%CI: 1.4, 2.3). compared to males^[6,10]^. Studies from China^[29]^, Easten Ethiopia^[30]^ and Ghana^[26]^ also supported this findings and reported that the prevalence of depression was higher in females than males. Although the reason for the higher prevalence of depression symptoms among women is not clearly understood. The possible reasons for this association may be that women are frequently expected to shoulder more caregiving and household responsibilities and suffer disproportionately from gender-based violence, such as sexual and domestic abuse, which raises their risk of depression. The hormonal fluctuations that occur during menstruation, pregnancy, the postpartum period, and menopause can increase the vulnerability of women to mood disorders and consequently to depression symptoms.

Exposure to secondhand smoking is found to be significantly associated with anxiety symptoms but not with depression or stress symptoms. A cross-sectional study ^[31]^ and systematic review^[32]^ has supported this findings. The possible reason for this association might be that longtime exposure to secondhand smoke can cause shortness of breath, and raise heart rate, potentially intensifying anxiety symptoms. This study showed that Dalit, Janajati, and Madhesi have a higher risk of having stress symptoms compared to Brahmin/Chhetri. A study conducted by Thagunna et al. and Kohrt et al. in Jumla, Nepal, supported this finding, indicting higher rates of mental health issues among lower minority groups compared to high castes.^[22,33]^ The possible reason for this association is that less privileged ethnic groups tend to have lower socioeconomic status, less access to a diverse range of dietary products, and lesser access to comprehensive nutrition information compared to their highly advantaged counterparts. As a result, less privileged ethnic groups had an increased risk of stress.

In contrast to previous studies^[14,15]^, smoking habits was found to be non-significant in this study. A possible reason for this negative association may be that few participants (16.9%) in this study were current smokers. Findings of previous studies showed that hypertension, and excess body fat were significantly associated with depression symptoms^[7,30,34,35]^, but these are not significant in this study. The possible reasons for this negative association may be due to variation in study population, study settings, study variables, sample size, and sampling techniques. Another reason could be that CVDs risk factors are categorized using these variables. Since these variables are already included, they are found to be non-significant.

However, this study has some limitations as well that must be mentioned. First, quite less frequency of participants with zero CVDs risk factors were tapped, which may be cautiously interpreted when comparing. Secondly, binary logistic regression might have given less robust findings (to some extent).

## Conclusion

The study revealed nearly half of the participants have each of depression, anxiety, and stress symptoms. Inadequate intake of fruits and vegetables was observed among almost all, whereas overweight and obese and excess body fat were found among more than one-fourth. In comparison to 0 risk factor, participants with 2 and ≥3 CVDs risk factors had a moderate-to-strong positive association with mental health status. Mental health screening is warranted among the people with two and above CVD risk factors. Further, a stronger design with an appropriate number of participants in the group–without CVDs risk factors is recommended before making a firm conclusion.

## Data Availability

All data produced in the present work are contained in the manuscript.

## Acknowledgments

We would like to acknowledge all participants who provided their valuable time for our study. We would like to express our gratitude to Mr. Basant Kumar Dhami and Mr. Ramesh Shrestha for their support in data analysis in our manuscript. Additionally, we are thankful to all those who provided us with direct and indirect support during our study.

## Author Contributions

**Conceptualization:** Ramesh Ojha, Chiranjivi Adhikari, Hari Prasad Kaphle, Dikshya Adhikari

**Data curation:** Ramesh Ojha

**Formal analysis:** Ramesh Ojha, Chiranjivi Adhikari, Dikshya Adhikari

**Funding acquisition:** Ramesh Ojha

**Investigation:** Ramesh Ojha, Chiranjivi Adhikari

**Methodolgy:** Ramesh Ojha, Chiranjivi Adhikari, Hari Prasad Kaphle, Dikshya Adhikari

**Project administration:** Ramesh Ojha, Chiranjivi Adhikari

**Resources:** Ramesh Ojha, Chiranjivi Adhikari, Hari Prasad Kaphle, Dikshya Adhikari

**Software:** Ramesh Ojha, Chiranjivi Adhikari, Dikshya Adhikari

**Supervision:** Chiranjivi Adhikari

**Validation:** Chiranjivi Adhikari

**Visualization:** Ramesh Ojha, Chiranjivi Adhikari

**Writing-original draft:** Ramesh Ojha, Chiranjivi Adhikari, Hari Prasad Kaphle, Dikshya Adhikari

**Writing-review & editing:** Ramesh Ojha, Chiranjivi Adhikari, Hari Prasad Kaphle, Dikshya Adhikari

